# Markers of psoriatic skin phenotype: androgen/estrogen and cortisone/cortisol imbalance, and DNA damage

**DOI:** 10.1101/2022.10.25.22281520

**Authors:** Şeyma Başar Kılıç, Serpil Taheri, Ecmel Mehmetbeyoğlu, Eda Öksüm Solak, Minoo Rassoulzadegan, Murat Borlu

## Abstract

Patients prone to psoriasis suffer after a breakdown of the epidermal barrier and develop poorly healing lesions with abnormal proliferation of keratinocytes.

Strong inflammatory reactions with genotoxicity (short telomeres) suggest impaired immune defences with DNA damage repair response (DDR) in patients with psoriasis. Recent evidence indicates the existence of crosstalk mechanisms linking the DDR machinery and hormonal signalling pathways that cooperate to influence both progressions of many diseases and responses to treatment.

The aim of this study was to clarify whether steroid biosynthesis and genomic stability markers are altered in parallel during the formation of psoriatic skin. Understanding between the interaction of the steroid pathway and DNA damage response is crucial to addressing underlying fundamental issues and managing resulting epidermal barrier disruption in psoriasis.

Twenty patients with psoriasis and fifteen healthy volunteers were included in this study. Transcription levels of estrogen (*ESR1, ESR2*), androgen (*AR*), glucocorticoid/mineralocorticoid receptors (*NR3C1, NR3C2*), *HSD11B1/HSD11B2* genes, and DNA damage sensors (*SMC1A, TREX1, TREX2, SSBP3, RAD1, RAD18, EXO1, POLH, HUS1*) were determined by Real-Time-PCR in blood and skin samples (Lesional, non-lesional) from psoriasis and control groups.

**Results:** We found that *ESR1, ESR2, HSD11B1, NR3C1, NR3C2, POLH*, and *SMC1A* transcripts were significantly decreased and *AR, TREX1, RAD1*, and *SSBP3* transcripts were increased dramatically in the lesional skin compared to skin samples of controls.

As a result, we found that the regulation of the steroidogenic pathway was disrupted in the lesional tissue of psoriasis patients and that a sufficient glucocorticoid and mineralocorticoid response did not form and the estrogen/androgen balance was altered in favour of androgens. We suggest that an increased androgen response in the presence of DDR increases the risk of developing psoriasis. Although this situation may be the cause or the consequence of a disruption of the epidermal barrier, our data suggest developing new therapeutic strategies.

## INTRODUCTION

Psoriasis is a chronic and recurrent inflammatory skin disease characterized by abnormal keratinocyte proliferation, vascular hyperplasia, and inflammatory cell infiltration in the dermis and epidermis. Disruption of the integrity or function of the epidermal barrier leads to epidermal hyperplasia and psoriasis [1,2]. It is generally accepted that the central pathogenesis of psoriasis is the dysfunction of T lymphocytes cells affected by complex interactions between genetic and environmental factors such as trauma, infection, stress, medications, smoking and alcohol consumption [3,4]. It has been revealed that TNF-alpha, interleukin 23, 17 and 22 play a major role, resulting from the main interaction of epidermal dendritic cells, T lymphocytes and keratinocyte cells. However, it is unclear what determines dendritic cell stimulation and T lymphocyte cells differentiation.

There are many complex immunological reactions in psoriasis resulting in epidermal hyper-proliferation with abnormal keratinocyte differentiation [5]. However, it is not yet known what triggers this mechanism, whether the inflammation that occurs with the breakdown of the epidermal barrier induces the proliferation of keratinocyte or the accumulated damage on the DNA of keratinocytes triggers the inflammation. Revealing which features of the epidermal barrier make up this cascade may explain parts of the disease that are still unclear.

Psoriasis has a family heritage, but so far no genes or mutations have been identified that could fully explain the mechanism of the disease. The disease affects both men and women. The skin is considered the extra-adrenal organ that plays an important role in the neuroendocrine-immune system by synthesizing a wide range of hormones from cholesterol to glucocorticoids and sex steroids [6]. It is important to maintain the regulation of lipids, cholesterol, ceramide, and epidermal DNA synthesis in the skin in order to preserve the integrity of the epidermal barrier [7–9].

The synthesis of steroid in the skin is very important for the maintenance of the epidermal barrier function. While a sufficient estrogen response ensures the moisture balance of the skin, testosterone plays the opposite role and its increase triggers the deterioration of the epidermal barrier [9–12]. Additionally, the balance of 11β-hydroxysteroid dehydrogenase type 1 (HSD11B1) and 11β-hydroxysteroid dehydrogenase type 2 (HSD11B2) enzymes, which convert active cortisol/inactive cortisone, is also very important for healthy skin. Increased or insufficient levels of glucocorticoid or their receptor responses (Glucocorticoid Receptor (NR3C1), Mineralocorticoid Receptor (NR3C2)) can also impair skin barrier function [8,13,14]. Disruption of epidermal barrier integrity and regulation of epidermal DNA synthesis also triggers the DNA damage response [7]. We have recently shown that the integrity of the genome is altered in patients predisposed to develop psoriasis [15].

We hypothesize that disruption of local regulation of steroidogenic activity is a threat to the integrity of the epidermal barrier that triggers DNA damage. The inability to sufficiently repair and the DNA fragments that occur during the repair of the damage trigger the immune system and cause inflammation. Finally, when DNA damage is not sufficiently repaired, cell proliferation altered. We sought to identify transcripts associated with disruption of the epidermal barrier, causing DNA damage and disrupting local steroidogenic activity in patients with psoriasis.

Revealing the features of the epidermal barrier that make up this cascade may explain parts of the disease that are unclear [16]. We determined the transcript levels of *estrogen* (*ESR1, ESR2*), *androgen* (*AR*), *glucocorticoid/mineralocorticoid receptors* (*NR3C1, NR3C2*), *HSD11B1/ HSD11B2* genes and *DNA damage sensors* (*SMC1A, TREX1, TREX2, SSBP3, RAD1, RAD18 EXO1, POLH, HUS1*) in the lesional and non-lesional skin tissue according to healthy skin. We reveal an imbalance of androgen/estrogen and cortisol/cortisone, as well as an increased response to DNA damage by a psoriatic skin phenotype.

## MATERIALS AND METHODS

This study was approved by the Human Ethics Committee of the Erciyes University on 20/02/2019, with the decision number 2019/121, and 20 patients who were diagnosed with chronic plaque-type psoriasis and admitted at the Erciyes University, Faculty of Medicine, Department of Dermatology and Venereal Diseases and age- and sex-matched 15 healthy volunteers without chronic disease were included after obtaining their consent. The patients and controls participating in the study were verbally informed and their written consent was obtained. Patients under the age of 18 and healthy controls were not included in the study. Psoriasis Area Severity Index (PASI) values were calculated [17] and recorded and patients with a PASI value greater than 10 were included in the study. Patients with an inflammatory/autoimmune or chronic disease other than psoriasis and patients who have received topical therapy for psoriasis within the last 2 weeks or systemic therapy in the last 4 weeks were not included in the study.

### Collection of skin biopsy and blood samples from psoriasis patients and healthy control groups

Skin samples were taken from the lesional skin and normal skin adjacent to the lesion from the 20 patients with psoriasis. Skin biopsy samples were taken from skin areas (popliteal fossa) located behind the knee of the healthy control group who accepted to participate in the study. When collecting skin biopsy samples, local anesthesia was applied under the skin, and tissue samples were taken with a 3 mm puncture tool after providing completely sterile conditions. In addition, 5 ml of peripheral venous blood was collected from patients and study control groups in a purple-capped EDTA tube. Tissue and blood samples were immediately delivered to the Genome and Stem Cell Center Genome Unit, RNA isolations were performed immediately, and RNA samples were stored at -80°C until at the start of the study.

### RNA isolation and Real -Time PCR

Total RNA was isolated from skin tissue and blood samples using TRIzol (Cat No: 11667165001 Roche, Germany and cDNA was synthesized from RNA samples obtained using a cDNA synthesis kit (Cat no:18091200, Thermo Fisher Scientific, USA). cDNA samples were quantified using the BioMark high-throughput Real-Time PCR system (Fluidigm, San Francisco, CA) for Q-PCR analysis at transcript levels [18,19]. We selected sixteen candidate transcripts known to be involved in **DNA damage and repair** ((*Structural Maintenance Of Chromosomes 1A* (*SMC1A*), *Three Prime Repair Exonuclease 1(TREX1), Three Prime Repair Exonuclease 2 (TREX2), Single Stranded DNA Binding Protein 3 (SSBP3), RAD1 Checkpoint DNA Exonuclease (RAD1), RAD 18 E3 Ubiquitin Protein Ligase (RAD18), Exonuclease 1 (EXO1), DNA Polymerase Eta (POLH)* and *Checkpoint Clamp Component (HUS1*)) and **local steroidogenic activity** ((*Estrogen Receptor 1(ESR1), Estrogen Receptor 2(ESR2), Androgen Receptor (AR), Glucocorticoid Receptor (NR3C1), Mineralocorticoid Receptor (NR3C2), Hydroxysteroid 11 Beta Dehydrogenase 1(HSD11B1), and Hydroxysteroid 11 Beta Dehydrogenase 2 (HSD11B2))* for determining transcript levels. Primer sequences were given in the supplementary Table 1. *GAPDH* and *ACTB* genes were used as housekeeping genes. All samples were analyzed in duplicate, and values were normalized using the 2^―ΔΔ*Ct*^ method [18–20].

### Statistical Analysis

Data were collected using BioMark Real-Time PCR Analysis Software, the linear derivative baseline correction method, and the auto global Ct threshold method. System-derived Ct values of 999 and values larger than 26 were considered non-specific and beyond detection limits and were therefore removed. Median Limit of detection (LOD)Ct values were calculated across all arrays to impute missing values [21]. Data normalization was performed using the 2^―ΔΔ*Ct*^ method [20]. The compliance of the data to the normal distribution was evaluated using a histogram, q-q graphs, and the Shapiro-Wilk test. Variance homogeneity was evaluated using the Levene test. The one-way ANOVA test was used to compare the differences between the 16 different transcripts levels of the lesional and non-lesional skin tissues obtained from the patient group and skin tissues and blood samples obtained from the healthy control group. The relationship between the quantitative data was evaluated using Spearman correlation analysis. Data were analyzed using Graphpad Prism (version 8.0.1) software. P values <5% were considered statistically significant.

## RESULTS

Twenty patients with chronic plaque psoriasis were included in the study. Of these patients, 10 (50%) were female patients. The control group consisted of 15 people, 8 (53.33%) healthy women and 7 (46.67%) healthy men. The mean ages of the patient and healthy control groups were 39.2 and 38.5, respectively. There is no statistically significant difference between the groups in terms of age or gender (p> 0.05). The mean disease duration in the patient group was 14.4 years. The mean PASI was 15.8. The demographic characteristics of the patient and the control group are presented in Table 1.

**Table 1.**
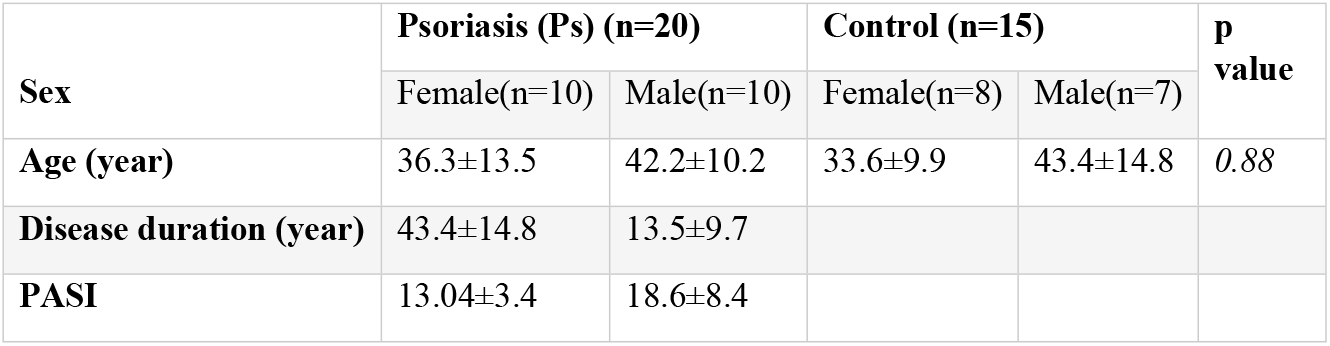
Demographic characteristics of the psoriasis patients and the control group.

| Sex | Psoriasis (Ps) (n=20) |  | Control (n=15) |  | p value |
| --- | --- | --- | --- | --- | --- |
|  | Female(n=10) | Male(n=10) | Female(n=8) | Male(n=7) |  |
| Age (year) | 36.3±13.5 | 42.2±10.2 | 33.6±9.9 | 43.4±14.8 | 0.88 |
| Disease duration (year) | 43.4±14.8 | 13.5±9.7 |  |  |  |
| PASI | 13.04±3.4 | 18.6±8.4 |  |  |  |

### Imbalance detected in the steroidogenic and DNA damage pathway in the lesional and non-lesional skin tissue of patients with psoriasis

Transcripts levels were determined in skin biopsy and blood samples by qRT-PCR, the results are shown in Figures 1, 2 and Tables 2, 3.

**Table 2.** Transcript profiles of the steroidogenic pathway, DNA damage and repair genes in the blood of psoriasis patients (Ps) and the healthy control group.

|  | Genes | Ps (n=20) | Control (n=20) | P value |
| --- | --- | --- | --- | --- |
| Steroidogenic Activity | *ESR1* | 0.9(0.7-1) | 1(0.7-1.2) | 0.420 |
|  | *ESR2* | 0.3(0.2-0.8) | 0.3(0.2-2.6) | 0.49 |
|  | *AR* | 0.6(0.3-1.1) | 1.1(0.6-1.9) | 0.055 |
|  | *NR3C1(GR)* | 0.6(0.3-1.1) | 0.5(1.2-5) | 0.828 |
|  | *NR3C2(MR)* | 0.2(0.1-0.7) | 1.3(0.2-17.8) | 0.0570 |
|  | *HSD11B1* | 1.2(0.9-1.4) | 1(0.3-4.9) | 0.75 |
|  | *HSD11B2* | 1.5(1.1-1.9) | 1.2(0.6-2) | 0.515 |
| DNA Damage Sensors and Repair | *RAD1* | 1(0.8-1.3) | 1.1(0.9-1.4) | 0.633 |
|  | *RAD18* | 0.2(0.1-0.8) | 0.9(0.1-2.6) | 0.171 |
|  | *SMC1A* | 0.8(0.2-1.9) | 1.6(0.2-8.1) | 0.419 |
|  | *EXO1* | 0.3(0.1-0.7) | 0.2(0.1-1.8) | 0.707 |
|  | *HUS1* | 0.7(0.5-1.1) | 0.7(0.2-2.9) | 0.495 |
|  | *TREX1* | 1.1(0.9-1.4) | 1.1(0.8-1.5) | 0.831 |
|  | *TREX2* | 1.8(1.3-2.2) | 1.2(0.8-1.4) | 0.0688 |
|  | *SSBP3* | 0.9(0.7-1) | 1(0.7-1.2) | 0.419 |
|  | *POLH* | 0.3(0.1-0.8) | 0.8(0.1-2.3) | 0.363 |
| The data are expressed as median (1st and 3rd quartile). Different letters show the difference between groups |  |  |  |  |

**Table 3.** The transcript profiles of the steroidogenic pathway, DNA damage and repair genes in the skin biopsies of psoriasis patients (Lesional, Non-lesional) and the healthy control group.

|  | Genes | Ps lesional tissue (n=20) | Ps non-lesional tissue (n=20) | Control tissue(n=15) | p value |
| --- | --- | --- | --- | --- | --- |
| Steroidogenic Pathway | *ESR1* | 0.3(0.1-0.7)<sup>a</sup> | 0.5(0.3-0.6)<sup>ab</sup> | 1(0.5-2.1)<sup>b</sup> | **0.0078** |
|  | *ESR2* | 0.4(0.1-0.9)<sup>a</sup> | 0.8(0.3-1.3)<sup>ab</sup> | 1.1(0.4-2.8)<sup>b</sup> | **0.0161** |
|  | *AR* | 2.2(0.9-7.6)<sup>a</sup> | 0.4(0.2-2)<sup>b</sup> | 0.5(0.3-1)<sup>b</sup> | **0.0049** |
|  | *NR3C1(GR)* | 0.5(0.2-1)<sup>a</sup> | 1.2(0.6-2)<sup>b</sup> | 1.2(0.7-1.7)<sup>b</sup> | **0.0132** |
|  | *NR3C2(MR)* | 0.2(0.04-0.3)<sup>a</sup> | 0.5(0.3-1.5)<sup>b</sup> | 0.8(0.5-2)<sup>b</sup> | **<0.0001** |
|  | *HSD11B1* | 0.04(0.01-0.1)<sup>a</sup> | 0.2(0.1-0.6)<sup>b</sup> | 0.7(0.3-1.3)<sup>b</sup> | **<0.0001** |
|  | *HSD11B2* | 2.1(0.6-4.3) | 1(0.7-2.3) | 0.8(0.6-1.7) | 0.3786 |
| DNA Damage Sensors and Repairs | *RAD1* | 3.7(1.9-10.3)<sup>a</sup> | 0.7(0.2-3.6)<sup>b</sup> | 0.8(0.3-2.2)<sup>b</sup> | **0.0013** |
|  | *RAD18* | 0.5(0.1-1.1) | 0.4(0.2-1.2) | 0.9(0.7-1.6) | 0.1349 |
|  | *SMC1A* | 0.5(0.2-0.8)<sup>a</sup> | 0.8(0.6-1.2)<sup>ab</sup> | 1.1(0.8-1.9)<sup>b</sup> | **0.008** |
|  | *EXO1* | 1.1(0.4-2.4) | 0.5(0.4-1.4) | 1(0.8-1.4) | 0.2002 |
|  | *HUS1* | 1(0.5-2.3) | 0.5(0.4-1.6) | 1.1(0.7-1.5) | 0.146 |
|  | *TREX1* | 2.2(1.5-5)<sup>a</sup> | 0.6(0.3-1.8)<sup>b</sup> | 0.7(0.4-1.6)<sup>b</sup> | **0.0067** |
|  | *TREX2* | 1.4(0.6-4) | 1.6(0.7-2.1) | 1.3(0.8-1.9) | 0.6423 |
|  | *SSBP3* | 3.4(1.4-6.6)<sup>a</sup> | 0.4(0.1-2.1)<sup>b</sup> | 0.6(0.2-2.2)<sup>b</sup> | **0.005** |
|  | *POLH* | 0.4(0.2-1)<sup>a</sup> | 0.8(0.4-1.8)<sup>ab</sup> | 1.2(0.8-1.4)<sup>b</sup> | **0.0208** |
|  | *The data are expressed as median (1st and 3rd quartile). Different letters show the difference between the groups.* |  |  |  |  |

**Figure 1.**
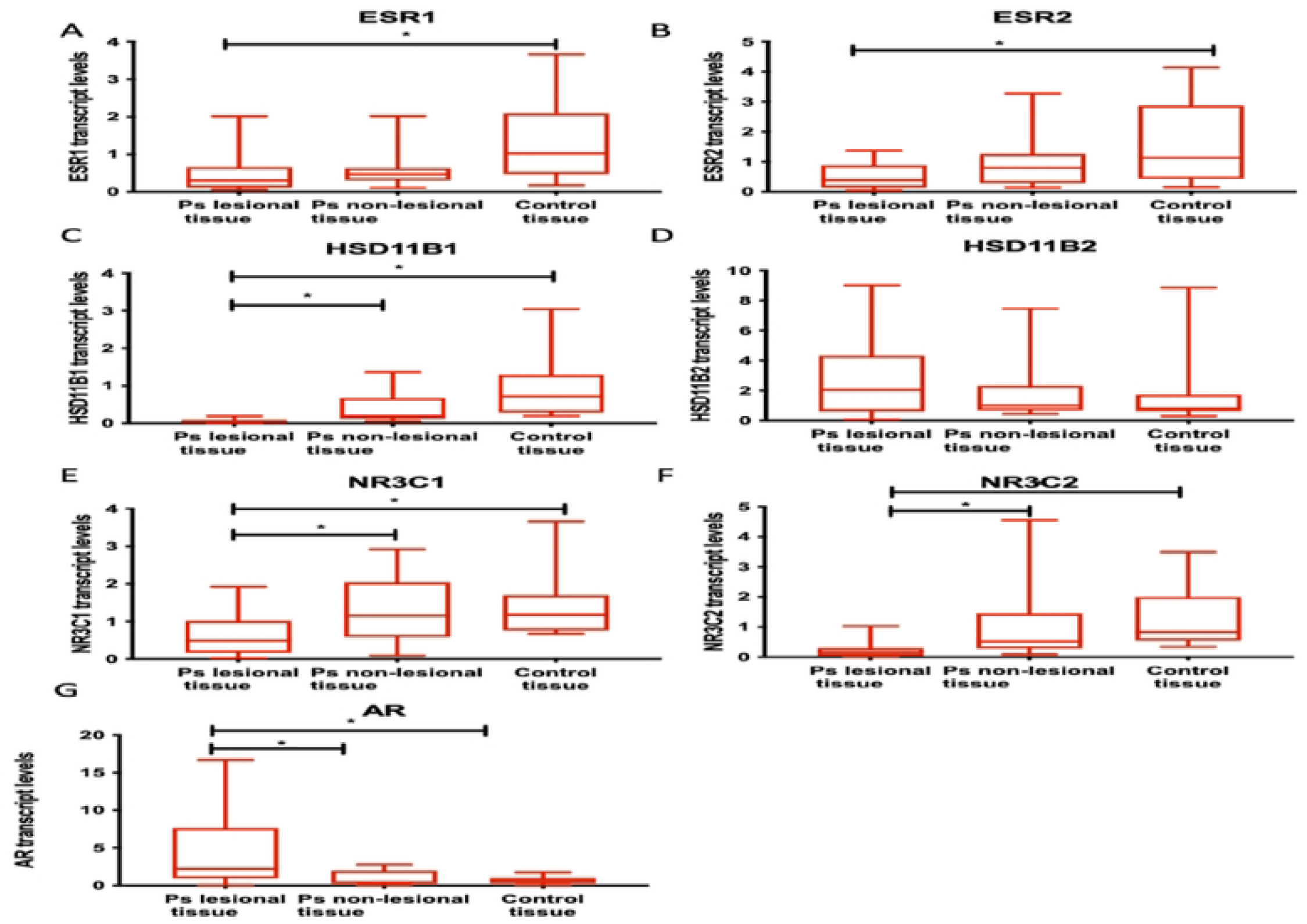
Transcription levels of genes involved in the local steroidogenic pathway in the lesional and non-lesional skin tissue samples from patients with psoriasis according to the control group. When the transcript levels of local steroidogenic pathway genes in the lesional and ton-lesional skin tissue samples from patients with psoriasis were compared to tissue samples from healthy volunteers, it was determined that the transcript levels of *ESR1 (****p: 0.0078)*** and *ESR2* ***(p: 0.0161)*** genes were significantly decreased in the lesional issue of patients with psoriasis compared to the control group. Transcription levels of *HSD11B1****(p <0.0001)***, *NR3C1* ***(p: 0.0132)***, and *NR3C2* ***(p <0.0001)*** were significantly decreased in the lesional tissue according to the non-lesional skin tissue and the control group. *AR* gene transcript levels were significantly increased in the lesional issue compared to non-lesional tissue and the control group ***(p: 0.0049)***. There was no statistically significant difference between groups in *HSD11B2* transcript levels ***(p: 1.3786)***.

**Figure 2.**
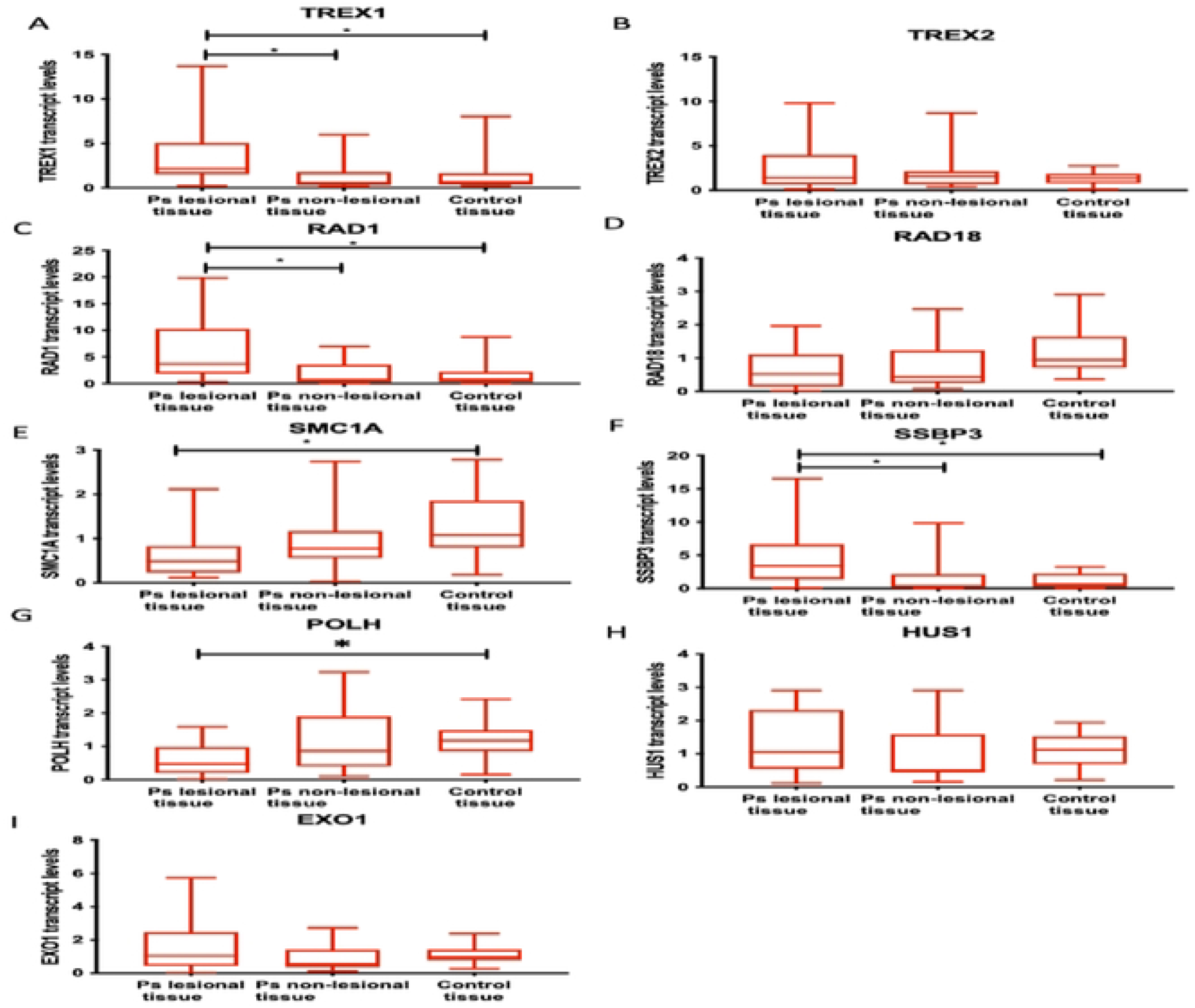
Transcription levels of DNA damage sensor and repairer genes in the lesional and non-lesional skin tissue samples from patients with psoriasis (Ps) according to control group. When the transcript levels of the DNA damage sensor and damage genes were compared from tissue samples taken from healthy volunteers and lesional and non-esional tissues from patients with psoriasis, the transcript levels of *TREXI (****p: 0.0067****), RAD1 (****p: 0.0013***), and *SSBP3 (****p: 0.005***) were significantly increased in the lesional issues compared to non-lesional tissues and control group. Transcription levels of *POLH* ***(p: 0.0208)*** and *SMC1A (****p: 0.008)*** genes were significantly decreased in the lesional tissue compared to tissue samples from the control group. There was no significant difference between the groups in the transcription levels of the *TREX2, RAD18, EXO1*, and *HUS1* genes.

The graphical summary of the heat map of transcripts in skin tissues is shown in Figure 3.

**Figure 3.**
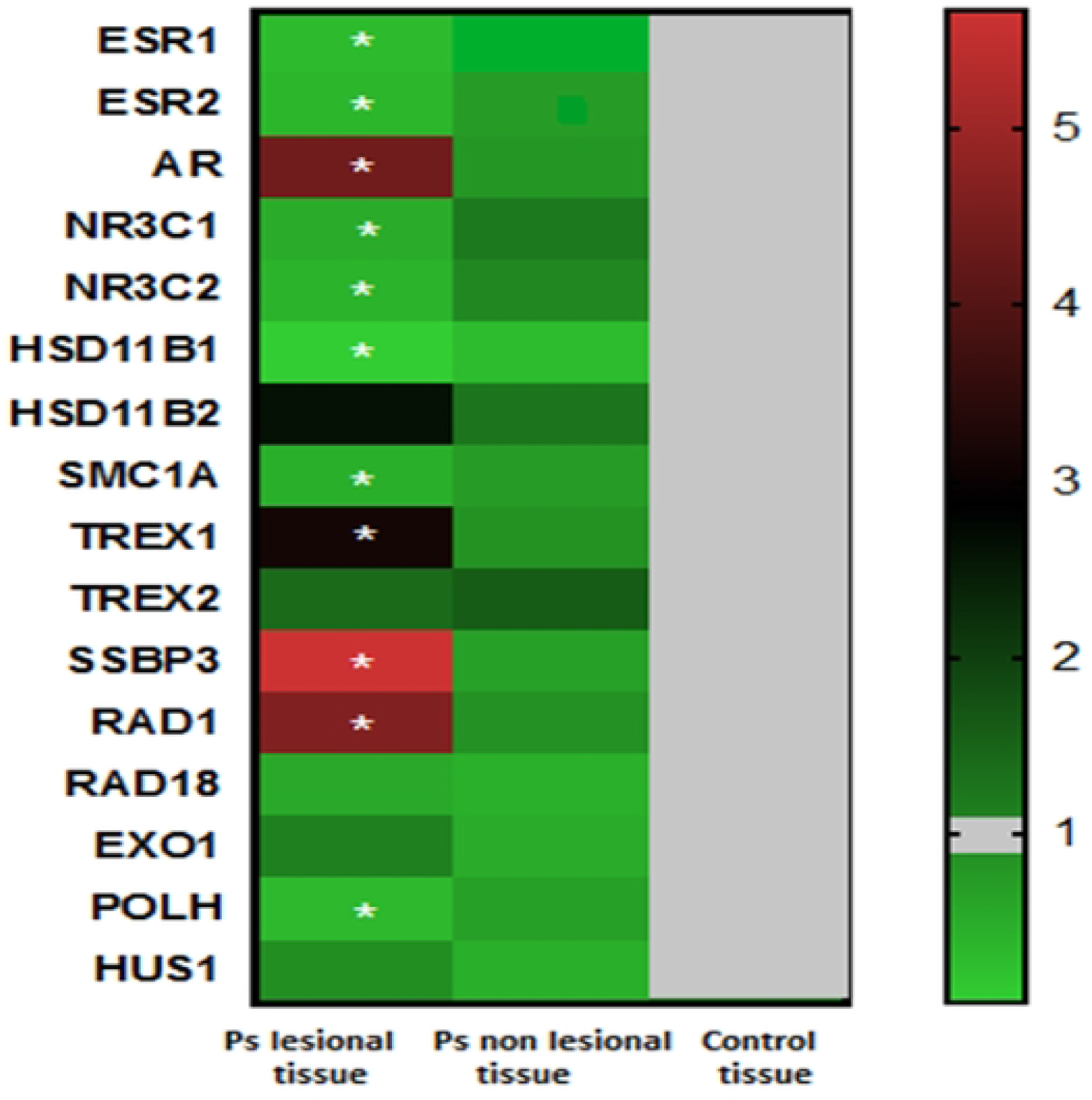
Heatmap graph of altered transcript profiles in skin biopsy samples from patients with psoriasis compared to control group (green color indicates genes with decreased transcript levels to control group, red color indicates genes with higher levels to control group). The transcription levels of *ESR1, ESR2, NR3C1, NR3C2, HSD11B1, SMC1A, POLH* genes were significantly decreased in the lesional tissue according to the control group, while the transcription levels of *AR, TREX1, SSBP3* genes were significantly increased in the lesional tissue according to the control group. There was no significant difference between the groups in the transcription levels of the *TREX2, RAD18, EXO1*, and *HUS1* genes.

Transcript levels of *ESR1 (Estrogen Receptor 1), ESR2 (Estrogen Receptor 2), HSD11B1(Hydroxysteroid 11 Beta Dehydrogenase), NR3C1(Glucocorticoid Receptor), NR3C2 (Mineralocorticoid Receptor), POLH (DNA Polymerase Eta)*, and *SMC1A (Structural Maintenance Of Chromosomes 1A)* in the lesional skin samples from psoriasis patients were significantly decreased compared to the control group, while transcription levels of *AR (Androgen Receptor), TREX1 (Three Prime Repair Exonuclease 1), RAD1 (RAD1 Checkpoint DNA Exonuclease)*, and *SSBP3 (Single-Stranded DNA Binding Protein 3)* were significantly increased in lesional skin samples of patients by compared to the control group see Figure 1and 2.

There was no significant difference between the groups in the transcript levels of *HSD11B2 (Hydroxysteroid 11 Beta Dehydrogenase 2), TREX2 (Three Prime Repair Exonuclease 2), RAD18 (RAD 18 E3 Ubiquitin Protein Ligase), EXO1(Exonuclease 1)*, and *HUS1(Checkpoint Clamp Component)* see Figure 1 and 2.

There was no difference in transcript levels in the non-lesional skin tissues and blood of patients compared to a control group. Blood transcript profiles of the healthy control group and patients with psoriasis are shown in Supplementary Table 2.

## DISCUSSION

We have recently shown that in patients prone to psoriasis, genome integrity is compromised with increased levels of TERRA and shortened telomere length in lesion tissue [22]. It is a cause or a result? We do not know yet. We investigated novel pathways in the development of psoriasis disease. The integrity of the epidermal barrier is very important for the regulation of epidermal DNA synthesis [23]. Impaired local steroidogenic activity function causes disruption of the epidermal barrier of the skin as well as inflammatory and autoimmune diseases [8]. Here, transcripts implicated in protecting epidermal barrier function and regulating epidermal DNA synthesis have been reported in the lesional and non-lesional psoriasis skin. Overall, our results reveal that the local steroidogenic pathway was increased with the androgen response, in contrast to the decreased estrogen response in psoriatic skin.

According to our results, the transcription levels of *Estrogen receptors (ESR1* and *ESR2)* were significantly decreased in the lesional skin samples compared to the control group and the transcription levels of *Androgen receptor* (*AR*) were significantly increased in the lesional skin samples compared to samples of non-lesional skin and the control group. This indicates an increase in androgen and a decrease in estrogen response in psoriatic skin tissue. In contrast to this, in blood samples, there was no difference between patient and control groups in *ESR1, ESR2*, and *AR* transcript levels. Accordingly, the increased androgenic response is observed locally only in psoriatic skin. Dermal effects of estrogen include improved development and function of the epidermal barrier and increased synthesis of collagen I and III in keratinocytes [17]. Unlike the effects of estrogen, androgens trigger epidermal hyperplasia and disrupt epidermal barrier function. Fluctuations in testosterone levels affect epidermal barrier balance and barrier function improves in the absence of androgens [8]. Similarly, in a study with pregnant mice, exogenous estrogen given to pregnant mice accelerated the development of the fetal epidermal barrier. In contrast, dihydro-testosterone delays the formation of the epidermal barrier [24]. Our results indicate that the androgen/estrogen balance in the skin of patients with psoriasis is impaired in the direction of increased androgen response and decreased estrogen response, and disruption of the epidermal barrier is triggered in psoriatic skin, according to the literature. Thus, changes in the hormonal balance of skin tissue have an etiological role in the development of psoriatic lesions and trigger hyper-proliferation. It is now desirable to consider new treatment options that could adjust the levels of androgen and estrogen and their receptors in skin tissues, which could improve skin recovery in patients.

Table 4 are summarized the function and the results shown here of the groups of transcripts. We report increased levels of three transcripts: *TREX1, RAD1* and *SSBP3* and decreased levels of *HSD11B1, POLH* and *SMC1A*, one of the largest cohesin subunits in lesion tissues.

**Table 4.** The function of the differentially expressed genes found to be associated with psoriatic skin phenotype

| Transcript Names | Gene Symbols | Chromosomal Localization | Gene Function | References |
| --- | --- | --- | --- | --- |
| *Estrogen Receptor 1* | *ESR1* | 6q25.1-q25.2 | It is the alpha receptor of estrogen that controls many cellular processes, including the growth, differentiation, and function of the reproductive system.<br>estrogen supports improved development and function of the epidermal barrier and increased synthesis of collagen. | [45][24][12][6][14] |
| *Estrogen Receptor 2* | *ESR2* | 14q23.2-q23.3 | It is the beta receptor of estrogen that controls many cellular processes, including the growth, differentiation, and function of the reproductive system.<br>Dermal effects of estrogen include improved development and function of the epidermal barrier and increased synthesis of collagen in skin | [45][24][12][6][14] |
| *Androgen Receptor* | *AR* | Xq12 | It is the receptor of androgens that triggers epidermal hyperplasia and disrupts the epidermal barrier function. | [45][9][12][6][24][14][46] |
| *Glucocorticoid Receptor* | *NR3C1* | 5q31.3 | Provide adequate glucocorticoid responses | [13][25][28][45][8][14][46] |
| *Mineralocorticoid Receptor* | *NR3C2* | 4q31 | Provide adequate mineralocorticoid responses | [13][26][25][45] |
| *Hydroxysteroid 11 Beta Dehydrogenase 1* | *HSD11B1* | 1q32.2 | Converts cortisone to cortisol in an active form | [45][13][8][14][26] |
| *Hydroxysteroid 11 Beta Dehydrogenase 2* | *HSD11B2* | 16q22.1 | Converts cortisol to cortisone in an inactive form | [45][13][8][14][26] |
| *Structural Maintenance Of Chromosomes 1A* | *SMC1A* | Xp11.22 | Encodes the largest structural subcomponent of cohesin, which ensures proper segregation and condensation of chromosomes. | [43][44][42][47] |
| *Three Prime Repair Exonuclease 1* | *TREX1* | 3p21.31 | 3'-5'exonuclease, responsible for DNA repair and, act as a DNA damage sensor, breaks cytoplasmic DNA into nucleotides to prevent the development of autoimmune responses | [34][30][29][48][32] |
| *Three Prime Repair Exonuclease 2* | *TREX2* | Xq28 | 3'-5'exonuclease, breaks down damaged nucleic acids and leads to activation of the inflammatory response. | [34][30][29][48][32] |
| *Single Stranded DNA Binding Protein 3* | *SSBP3* | 1p32.3 | Binds to replication fork strands and prevents the reformation of hydrogen bonds between the strands for DNA synthesis to function properly.<br>Increased levels are a sign of increased DNA replication and DNA damage repair. Related to the hydration of the skin, its expression increases in the differentiation process of keratinocytes. | [41] |
| *RAD1 Checkpoint DNA Exonuclease* | *RAD1* | 5p13.2 | DNA damage sensor,<br>The post-replicative enzyme that intervenes after DNA replication, encodes a component of a heterotrimeric cell cycle checkpoint complex that is activated to halt cell cycle progression in response to | [35][33][49] |
|  |  |  | DNA damage. |  |
| ***RAD 18 E3 Ubiquitin Protein Ligase*** | ***RAD18*** | 3p25.3 | DNA damage sensor, Required for the post-replication repair of damaged DNA. | [50][40][38][44] |
| ***Exonuclease 1*** | ***EXO1*** | 1q43 | Encodes a protein with 5' to 3' exonuclease activity as well as an RNase H activity, Involved in mismatch repair and recombination. | [40][47][33][38][51] |
| ***DNA Polymerase Eta*** | ***POLH*** | 6p21.1 | Involved in DNA repair and lesional DNA synthesis, the indicator of increased DNA damage and repair, | [39][52] |
| ***Checkpoint Clamp Component*** | ***HUS1*** | 7p12.3 | DNA damage sensor, thought to be an early DNA damage checkpoint signal molecule. | [35][40][34] |

These transcripts are involved in the various cellular processes and are listed below:

Local production of glucocorticoids occurs in different tissues, including lymphoid, brain and skin. Recent studies have highlighted the importance of local glucocorticoid biosynthesis in maintaining skin homeostasis [13,25]. While Hydroxysteroid 11-Beta Dehydrogenase-1(HSD11B1) converts cortisone to its active form, cortisol, Hydroxysteroid 11-Beta Dehydrogenase-2 (HSD11B2) converts cortisol to cortisone in an inactive form [26]. In our study, we found that *HSD11B1* transcript levels decreased in the lesional skin tissue of psoriasis patients compared to non-lesional and control group skin tissue, and there was no change in *HSD11B2* transcript levels. In one study, in vitro inhibition of *HSD11B1* transcripts was shown to increase keratinocyte and fibroblast proliferation [27]. Thus, the decrease in *HSD11B1* transcript levels in the lesional skin of patients with psoriasis may be an indication that cortisol levels necessary to suppress inflammation and maintain skin homeostasis are not being produced. It can also support the literature on this topic by explaining the possible cause of keratinocyte hyper-proliferation in lesional tissue [8,27].

The glucocorticoid receptor (GR, NR3C1) and mineralocorticoid receptor (MR, NR3C2), which provide adequate glucocorticoid and mineralocorticoid responses in tissue, are members of the nuclear receptor superfamily. The roles of these receptors in inflammatory skin diseases are currently being studied to develop new treatment options. Studies show that a combined loss of epidermal *GR* and *MR* genes in knockout mouse models leads to acute inflammation and psoriasis [13]. In a mouse study, impaired epidermal barrier, epidermal hyperplasia, hyperkeratosis, parakeratosis, and abnormal keratinocyte differentiation were found in keratinocyte-specific *GR* gene knockout mice. In recent years, unlike the effects of MRs on other organs, its effects on the skin would be similar to those of GRs [28]. According to our results, this could be due to the decrease in the transcript levels of the *NR3C1* and *NR3C2* genes in the lesional tissue of the patients. At these levels glucocorticoids and mineralocorticoids could not create an adequate response in the lesional tissues, therefore, inflammation could not be suppressed and abnormal hyper proliferation occurred in psoriatic skin.

The Three Prime Repair Exonuclease enzymes (TREX1 and TREX2) are exonucleases responsible for DNA repair and degradation and act as DNA damage sensors. TREX1 separates cytoplasmic DNA into nucleotides that are not detected by Pattern Recognition Receptors (PRPs) to prevent the development of type 1 interferon (IFN-1) responses and thus autoimmune responses [29]. The enzyme TREX2 is also a 3*’*-5*’*exonuclease similar to TREX1. However, it is expressed in tissues such as skin, tongue, and esophagus. TREX2 breaks down damaged nucleic acids and leads to activation of the inflammatory response [30]. Li et al. reported that TREX2 transcript levels were significantly increased in psoriatic skin [31]. Manilis et al. found that loss of TREX2 eliminates inflammation in two psoriasis murine models [32]. Here, our results reveal increased levels of *TREX1* transcript in the lesional skin tissue compared to non-lesional tissue and the control group, whereas there was no difference in *TREX2* transcript levels between the groups.

RAD1 Checkpoint DNA Exonuclease (RAD1), and RAD 18 E3 Ubiquitin Protein Ligase (RAD18) are DNA damage sensors. RAD18 is a post-replicative enzyme that intervenes after DNA replication. The *RAD1* gene encodes a component of a heterotrimeric cell cycle checkpoint complex that is activated to halt cell cycle progression in response to DNA damage [33]. An increase in *RAD1* transcripts indicates DNA damage and a decrease leads to insufficient repair [34]. Here, we found that the *RAD1* transcript levels increased in the lesional skin tissue compared to non-lesional tissue and the control group, and there was no difference between groups in *RAD18* transcript levels. The increase in *RAD1* transcripts in the lesional tissue is an indicator of DNA damage in the skin cells of patients with psoriasis. Han et al. found that keratinocytes isolated from *RAD1* knockout mice showed significantly more DNA double breaks and that these keratinocytes proliferated more slowly, and they suggested that this situation increased the susceptibility to the development of skin tumor [35]. Consistent with the literature, our results indicate the role of DNA damage and *RAD1* transcripts in the emergence of psoriatic skin phenotype.

Checkpoint Clamp Component (HUS1) and Exonuclease 1 (EXO1) are sensors responsible for DNA damage repair. EXO1 is a 5*’*nuclease. In a study by Hibner et al. in 2014, they evaluated the effect of Cyclosporin A (CsA) on normal human dermal fibroblasts through DNA repair genes. They found that CsA suppresses the *EXO1* transcripts. In this case, they suggested that CsA could cause tumor development by suppressing DNA repair genes [36].

CsA causes inhibition of cellular and humoral immune responses and has been a calcineurin inhibitor that has been used for many years in the treatment of severe psoriasis cases [37]. Patients who did not receive systemic treatment within the last four weeks were included in our study, and we found that there was no difference in the levels of *EXO1* and *HUS1* genes transcription in the tissues and blood samples from patients with psoriasis compared to the control group. Our results show that the *HUS1* and *EXO1* transcripts play no role in the formation of the psoriatic phenotype.

Eta DNA Polymerase (POLH) is a polymerase that is involved in DNA repair and lesion DNA synthesis. Increased *POLH* transcript levels are an indicator of increased DNA damage and repair, and its transcripts decreased are an indicator of insufficient repair [38]. In a study by Flanagan et al, it was found that although there was no *POLH* gene mutation in skin biopsies taken from patients diagnosed with basal and squamous cell carcinoma, transcript levels were decreased by compared to control skin tissue. We found that *POLH* transcript levels were decreased in the lesional skin tissue, consistent with data from Flanagan et al. This result indicates DNA damage and the role of *POLH* in psoriatic skin phenotype [39].

Single Stranded DNA Binding Protein 3 (SSBP3) is a protein that binds to replication fork strands and prevents the reformation of hydrogen bonds between the strands for DNA synthesis to function properly. Increased SSBP3 levels are a sign of increased DNA replication and DNA damage repair in any tissue [40]. In addition, SSBP3 is related to hydration of the skin, and its expression increases with the differentiation of keratinocyte [41]. Here our results indicate that *SSBP3* transcripts increased in the lesional skin compared to non-lesional skin and the control group. Our data suggest that *SSBP3* is associated with both increased DNA synthesis and damage due to excessive keratinocyte proliferation.

Cohesin is a DNA repair sensor which ensures the condensation of chromosomes during mitosis and meiosis, and plays a critical role during DNA synthesis and in particular in the repair of double-stranded breaks in the DNA. Structural Maintenance of Chromosomes 1A (SMC1A) is a protein that ensures proper separation of chromosomes and encodes the structural component of cohesion [42]. Decreased levels of cohesion lead to heterochromatinization and decreased expression of genes that act as regulators for cell regeneration, thereby dysregulating epidermal differentiation [43,44]. However, the relationship between cohesin and skin diseases and its mode of expression in tissues have not yet been revealed. According to our results, transcript levels of *SMC1A*, one of the largest cohesin subunits, were decreased in the lesional skin tissue of patients with psoriasis leading to increased DNA damage and heterochromatinization and dysregulation of keratinocyte differentiation in psoriatic skin.

## Conclusion

Steroidogenic pathways are perturbed in the lesional tissue of patients with psoriasis and the damaged DNA cannot be repaired adequately. A higher androgen response triggers disruption of the epidermal barrier and perturb genome integrity due to increased DNA damage. We have previously shown that genome integrity is impaired in psoriasis [22] and suggest that this triggers the breakdown of the epidermal barrier and the development of lesions in patients with psoriasis.

**Table S1.**
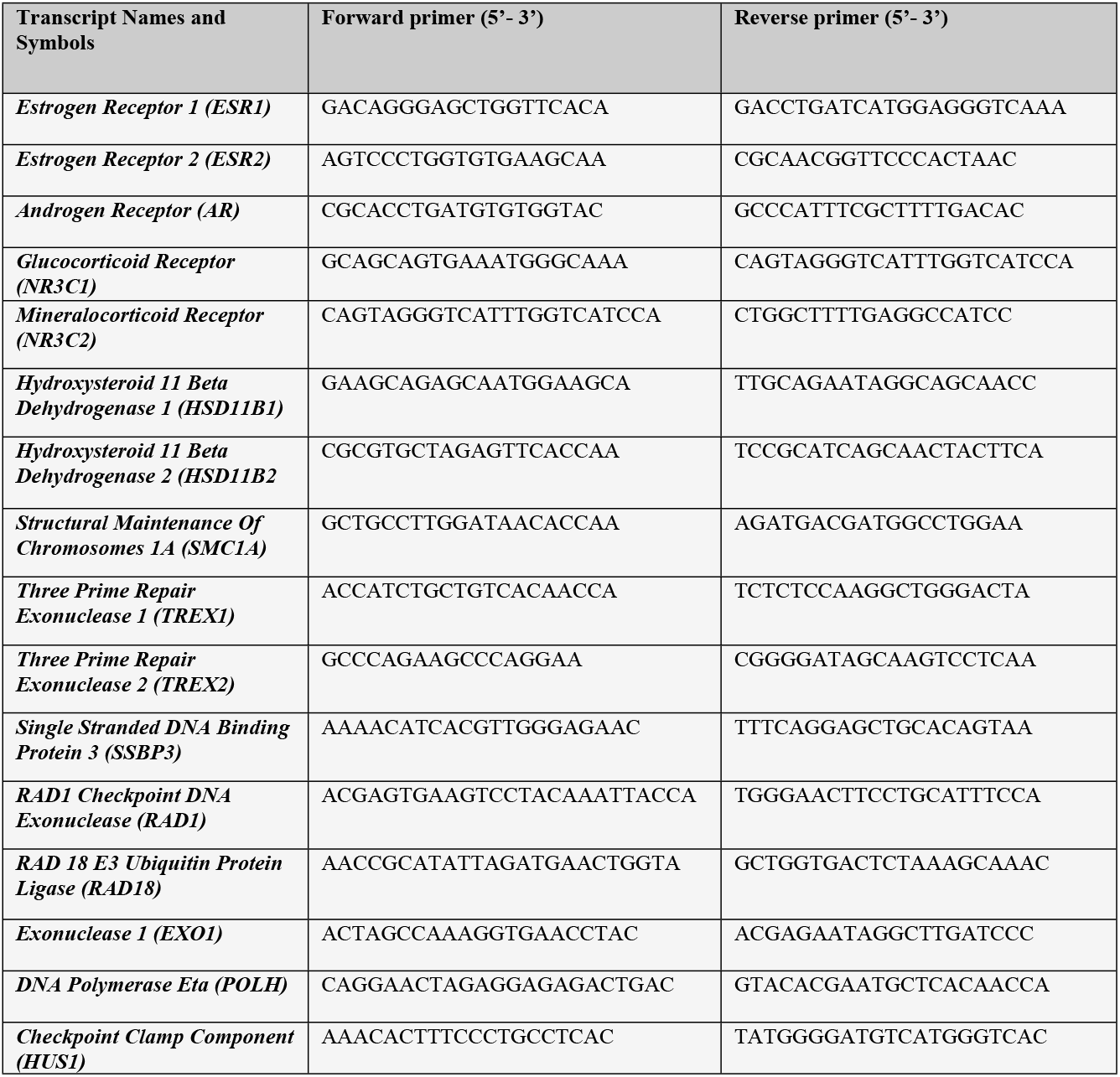
The list of primer sequences used for in this study

## Data Availability

All relevant data are within the manuscript and its Supporting Information files.

## Notes

**Financial support:** Supported by the Erciyes University Research Projects Unit. Project no: TCD-2019-9240

### Competing Interest Statement

The authors have declared that no competing interests exist.

### Funding Statement

Supported by the Erciyes University Research Projects Unit. Grant owner: Murat Borlu, Project no: TCD-2019-9240 https://bapsis.erciyes.edu.tr/Default2.aspx

### Author Declarations

This study was approved by the Human Ethics Committee of the Erciyes University on 20/02/2019, with the decision number 2019/121

